# Evaluating Goodness of Pronunciation and Phonological Posteriors as Objective Markers of Speech Severity in Motor Speech Disorders

**DOI:** 10.64898/2026.07.14.26358076

**Authors:** Fenqi Wang, Rene L. Utianski, Joseph R. Duffy, Leland R. Barnard, Hugo Botha

**Author notes:** (Corresponding author: Hugo Botha). The datasets of the current study are available from the corresponding author on reasonable request.

## Abstract

This study examined the extent to which goodness of pronunciation (GoP) scores and phonological posterior probabilities capture perceptual ratings of speech severity in individuals with motor speech disorders (MSD). Speech recordings of the word catastrophe were obtained from 489 participants, including 333 neurologically typical controls and 156 individuals with MSD. GoP scores were derived using traditional acoustic features and self-supervised speech representations, including WavLM and XLS-R, across multiple modeling approaches, while phonological posterior probabilities were extracted using Phonet. Model performance was evaluated using Kendall’s rank correlations, regression, and receiver operating characteristic analyses against speech-language pathologists’ perceptual ratings of sound distortion and intelligibility. Both GoP and phonological posterior probabilities were significantly associated with perceptual ratings. Self-supervised speech representations substantially outperformed traditional acoustic features, with WavLM-based GoP using k-nearest neighbors achieving the strongest performance. Across correlation, regression, and classification analyses, GoP consistently outperformed phonological posterior probabilities for both sound distortion and intelligibility. Age and gender had minimal influence on model-derived measures or their relationships with perceptual ratings. These findings demonstrate the value of self-supervised GoP as an objective measure of speech impairment while highlighting the complementary role of phonological posterior probabilities in characterizing articulatory aspects of motor speech disorders.

## I. Introduction

**M**OTOR speech disorders (MSDs), including dysarthria and apraxia of speech, are common consequences of neurological disease and reflect impairments in the planning, coordination, and execution of speech movements [1]. These disorders often lead to reduced articulatory precision and diminished intelligibility, with substantial impact on communication and quality of life [2]. Because specific patterns of speech impairment are associated with different neural substrates, careful characterization of speech can provide valuable clinical information for diagnosis, prognosis, and monitoring [1], [3]. However, as with many aspects of neurological assessment, accurate characterization of speech impairment remains challenging.

Auditory-perceptual evaluation is the current clinical standard for assessing MSDs, relying on expert judgment to rate features such as sound distortion and intelligibility [4]. While clinically meaningful, these assessments are inherently subjective, require specialized expertise, and are not widely accessible outside tertiary care settings [5], [6]. In addition, perceptual ratings often lack fine-grained resolution and may lack sensitivity to subtle or early changes in speech [7]. These limitations have motivated increasing interest in automated approaches that provide objective, scalable, and reproducible measures of speech impairment [8], [9].

### A. Automated Approaches to Speech Assessment

One widely used approach to quantifying segmental speech accuracy is Goodness of Pronunciation (GoP) [10], which evaluates how closely an acoustic realization matches its intended phoneme. Typically derived from acoustic model posteriors, GoP provides a continuous measure of phonetic deviation and has been applied in both second-language learning and clinical speech analysis [10], [11]. Prior work suggests that GoP is sensitive to articulatory abnormalities in disordered speech and can reflect perceptual dimensions of speech impairment [12], [13]. However, its relative effectiveness compared to alternative representations, particularly in capturing clinically relevant perceptual ratings, remains unclear. In addition, conventional GoP methods typically assume that phoneme distributions in the target population are relatively stable and that the training data sufficiently capture the range of speech patterns expected at test time. These assumptions may not hold in MSD populations, where speech production is often highly variable and atypical speech patterns are frequently underrepresented in training datasets [13], [14].

Recent advances in self-supervised speech models (S3Ms), such as WavLM [15] and XLS-R [16], offer a powerful alternative for representing speech. These models learn hierarchical representations from large-scale unlabeled data and have demonstrated strong performance across a range of speech processing tasks, including pronunciation assessment [17], [18]. By capturing both low-level acoustic structure and higher-level contextual information, S3M-derived features may better accommodate the variability and gradient distortions characteristic of MSD speech [12].

An alternative approach is provided by phonological posterior probabilities [19], which represent speech in terms of articulatory-phonological attributes rather than discrete phoneme categories. These features provide a linguistically grounded and interpretable representation of speech production and have been shown to differentiate disordered from typical speech and may reflect gradations of articulatory impairment [19], [20].

Although both GoP and phonological posterior approaches aim to quantify speech production at the segmental level, they differ in their underlying assumptions and representations. GoP evaluates the likelihood of a target phoneme given the observed acoustics, effectively measuring deviation from canonical pronunciation within a phoneme classification framework [10], [11]. In contrast, phonological posterior probabilities decompose speech into articulatory attributes (i.e., place and manner of production), providing a more direct representation of how speech is produced rather than how it is categorized [19]. As a result, GoP offers a compact, performance-oriented metric that aligns closely with recognition-based systems, whereas phonological posteriors emphasize interpretability and articulatory transparency. These differences suggest that the two approaches may capture partially overlapping but distinct aspects of speech impairment, with potentially differing sensitivity to perceptual severity.

### B. The Current Study

Despite these advances, several important gaps remain. First, although prior work on GoP has largely focused on applications such as second-language pronunciation assessment and non-native speech [10], [11], the impact of self-supervised representations on GoP performance in MSD populations is not well understood. Second, while phonological posterior probabilities offer interpretability [19], [20], their relationship to GoP in predicting clinically relevant judgments, such as sound distortion and intelligibility, has not been directly examined. Third, demographic factors such as age and gender, which are known to influence acoustic characteristics and, in some cases, speech intelligibility [21], [22], are rarely considered in these modeling frameworks.

The present study addresses these gaps by comparing several GoP computing approaches, with particular focus on how feature representations interact with those approaches, alongside phonological posterior probabilities obtained from Phonet. Using a large cohort of individuals with MSD and neurologically typical controls, we evaluate the extent to which these approaches capture perceptual ratings of sound distortion and intelligibility, and whether they provide overlapping or complementary information. Specifically, we ask:

1. Do self-supervised representations improve GoP-based assessment relative to traditional acoustic features?
2. To what extent do GoP and phonological posterior probabilities each relate to perceptual ratings of speech impairment, and do they provide redundant or partially complementary information?
3. Do age and gender influence model-derived measures or their relationships with perceptual severity?

From a speech processing perspective, this study examines how representation choice modulates the effectiveness of articulation, or pronunciation, assessment frameworks when applied to highly variable speech signals, such as those produced by speakers with MSD. By comparing traditional acoustic features with self-supervised representations across multiple GoP formulations and phonological posterior probability, this work isolates the contribution of representation learning to the accuracy and clinical relevance of automatic pronunciation assessment.

## II. Methods

### A. Dataset

The dataset included speech recordings of three repetitions of the word *catastrophe* from 539 participants, comprising 338 neurologically typical controls and 201 individuals with MSDs. Recordings with word error rate (WER) ≥ 1 were excluded to remove off-target productions. WER was determined by comparing the transcription generated by the Whisper ASR model (small) with the expected target word(s). This resulted in the removal of 5 controls and 45 individuals with MSDs. A sensitivity analysis conducted without WER-based filtering yielded the same overall patterns (see Supplemental material). Within the MSD group, participants were categorized by subtype as follows: apraxia of speech (AOS), phonetic subtype (*n* = 6), AOS, prosodic subtype (*n* = 13), ataxic dysarthria (*n* = 32), flaccid dysarthria (*n* = 26), hyperkinetic dysarthria (*n* = 46), hypokinetic dysarthria (*n* = 30), and spastic dysarthria (*n* = 25) [1]. The total across subtypes exceeds the number of unique MSD participants because 40 individuals presented with two co-occurring subtypes and one individual with three. In addition, a separate dataset consisting of 962 recordings from 904 healthy individuals without MSDs or neurological disease was used for model training. All participants provided written informed consent, and the study was approved by the Mayo Clinic Institutional Review Board.

### B. Goodness of Pronunciation

GoP was used to derive phoneme-level measures of articulatory accuracy. GoP quantifies the extent to which an observed acoustic segment corresponds to its intended phoneme and is commonly defined as the log posterior probability of a phoneme given the segment. Lower GoP values indicate greater deviation from expected phonetic realizations, making the measure sensitive to both categorical errors and gradient distortions in speech production. We computed GoP values by closely following the procedures in Choi, et al. [12], with only minimal customization, using their original code released on GitHub^1^.

#### 1) Feature Extraction and Segmentation

Acoustic representations were extracted using two types of feature encoders to capture complementary information about the speech signal.

Traditional acoustic features included Mel-frequency cepstral coefficients (MFCCs) and Mel spectrograms [23], which provide compact representations of spectral characteristics and have been widely used in pronunciation assessment. Self-supervised speech model (S3M) features were extracted from pretrained models, including WavLM [15] and XLS-R [16], which provide contextualized representations learned from large-scale unlabeled speech data. Features were extracted from multiple model layers to capture different levels of phonetic and contextual information [12].

For each utterance, phoneme-level segmentation was obtained using Montreal forced aligner [24], yielding start and end boundaries for each phoneme. Feature sequences were then partitioned according to these boundaries, and a single fixed-dimensional representation was derived per segment using temporal pooling. Center-based pooling was used to reduce the influence of boundary imprecision and coarticulatory transitions. This procedure resulted in a sequence of phoneme-aligned feature vectors for each utterance, which served as input for GoP computation.

#### 2) GoP Computing Approaches

We implemented two classes of approaches to compute GoP scores. Phoneme classifier-based methods estimate posterior probabilities over phoneme classes and compute GoP as the log posterior of the target phoneme (Eq. 1) [12]:

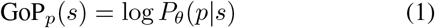

where *s* refers to a speech segment corresponding to a target phoneme *p*, and *P*_*θ*_(*p*|*s*) represents the estimated probability of producing phoneme *p* given the acoustic input *s*, computed by a model with learned parameters *θ*. These approaches rely on softmax normalization and assume a unimodal representation of each phoneme in the feature space. The following models were evaluated: Gaussian Mixture Model (GMM) [10], Neural Network (NN) [11], Deep Neural Network (DNN) [11], and MaxLogit [13]. Although these methods share a common classification framework, they differ in how pronunciation scores are derived from model outputs and how uncertainty is represented.

In contrast, out-of-distribution (OOD) detector-based methods quantify the degree to which an observed segment deviates from distributions learned from typical speech. In this approach, atypical pronunciations are treated as outliers relative to expected acoustic patterns, and GoP is derived from likelihood-or distance-based measures rather than posterior probabilities. The evaluated methods include k-nearest neighbors (kNN) [25], one-class support vector machine (oSVM) [26], phoneme-specific one-class SVM (p-oSVM) [27], and mixture-based GoP (MixGoP) [12]. MixGoP addresses the unimodal limitation by modeling each phoneme as a Gaussian mixture distribution, where the phoneme likelihood and corresponding score are defined as follows:

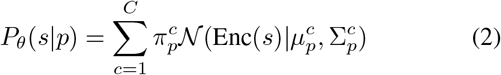

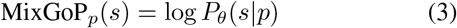

where *C* denotes the total number of Gaussian components used to represent each phoneme, 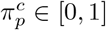 is the weight assigned to the *c*-th component for phoneme *p*, and 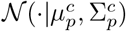 is a Gaussian distribution characterized by its centroid 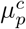 and spread 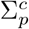. The term Enc(*s*) refers to the acoustic feature vector extracted from the speech segment *s* by a feature encoder. These approaches address the limitation that disordered speech may not be well represented within the training distribution and provide an alternative characterization of pronunciation variability.

#### 3) Evaluation

Phoneme-level GoP scores were aggregated to the utterance level by averaging across all phoneme segments within each utterance (i.e., recording) [12], as given by:

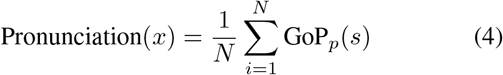

where *x* represents a full utterance comprising *N* phoneme segments, and GoP_*p*_(*s*) is the pronunciation score assigned to each segment *s* associated with its corresponding target phoneme *p*. This yielded a single GoP score per utterance, reflecting overall articulatory performance while preserving sensitivity to segment-level variation. Model performance was evaluated using the Kendall rank correlation coefficient (Kendall’s *τ*) between GoP scores and perceptual ratings of sound distortion and intelligibility provided by two board-certified speech-language pathologists [13]. Kendall’s *τ* was selected due to its robustness to non-normal distributions and its suitability for assessing monotonic relationships with ordinal severity metrics.

### C. Phonet

To complement and validate the GoP-derived measures, phonological posterior probabilities were extracted using Phonet [19], a neural network model that computes phonological posterior probabilities directly from continuous speech, providing an independent and articulatorily interpretable representation of speech production. Additional implementation details are available in the original GitHub repository^2^. While GoP quantifies deviations from expected phoneme realizations based on acoustic likelihoods, Phonet offers a feature-based characterization grounded in phonological structure, allowing for comparison across modeling approaches.

#### 1) Preprocessing and Feature Extraction

Although originally developed for Spanish, the model was retrained on the 360-hour LibriSpeech corpus using an English-specific phonological inventory to ensure compatibility with the present dataset. Audio recordings were preprocessed using standard procedures, including silence removal, resampling to 16 kHz, and amplitude normalization. Phoneme boundaries were obtained using the Montreal Forced Aligner [24], and a subset of alignments was manually reviewed to verify segmentation accuracy.

Phonet processes speech in overlapping frames and outputs posterior probabilities over 23 phonological classes. These probabilities reflect the likelihood that a given frame exhibits specific phonological attributes and can be interpreted as continuous measures of articulatory realization.

#### 2) Feature Aggregation and Selection

Segment-level features were computed by averaging posterior probabilities across frames within each phoneme. To reduce the influence of transitional regions and potential alignment inaccuracies, only the central portion of each segment was considered [28], with the number of contributing frames scaled according to segment duration. This approach emphasizes the most stable acoustic region of each phoneme while minimizing boundary-related variability, which may be more pronounced in speakers with MSD.

Following segment-level aggregation, posterior values were summarized according to broad phoneme classes. Specifically, consonantal posterior probabilities were used for consonants, and syllabic posterior probabilities were used for vowels. This grouping reflects primary articulatory distinctions, as consonants and vowels often exhibit different patterns in disordered speech [9].

### D. Clinical Assessment and Severity Measures

Perceptual judgments regarding the presence and classification of MSDs were conducted by two board-certified speech-language pathologists with expertise in neurogenic communication disorders. Evaluations were based on a brief battery of speech-language tasks, during which the clinicians provided detailed annotations of speech characteristics across tasks. These annotations were subsequently used to inform severity-related analyses.

Severity was characterized using two complementary perceptual ratings provided for each recording: sound distortion and intelligibility [29]. Sound distortion captures articulatory imprecision, reflecting deviations in the production of consonants, vowels, or both. Intelligibility reflects the overall clarity of speech and the extent to which spoken output can be understood by a blinded listener. Together, these measures provide complementary perspectives on speech impairment, with sound distortion indexing segmental accuracy and intelligibility capturing functional communication impact.

Sound distortion was rated on a six-point ordinal scale. Among the 156 participants with MSD, ratings were distributed as follows: 26 normal, 31 equivocal, 63 mild, 24 moderate, 2 marked. Two participants with severe sound distortion were excluded due to WER-based filtering. Intelligibility was rated on a three-point ordinal scale, with the following distribution in the MSD group: 121 normal, 18 equivocally reduced, and 17 abnormal.

### E. ROC Analysis

Receiver operating characteristic (ROC) analyses were conducted to evaluate the discriminative ability of two metrics, z-transformed GoP scores and log-odds posterior probabilities, using the pROC package in R [30]. Fifty controls were added to the MSD group and coded as absent for sound distortion and normal for intelligibility, allowing inclusion in both comparisons. Four ROC curves were computed by crossing the two metrics with two binary outcomes: presence versus absence of sound distortion and normal versus abnormal intelligibility. Predictor direction was specified such that lower z-scores corresponded to the clinical group. The area under the curve (AUC) was used to quantify classification performance, and optimal thresholds were determined using the Youden index.

Sensitivity and specificity analyses were conducted to evaluate clinical utility based on these thresholds. Participants were classified as positive if their values fell below the optimal threshold for each metric. In addition to evaluating each metric separately, two combined decision rules were examined: an OR condition, in which either metric met the threshold, and an AND condition, in which both metrics were required. Sensitivity and specificity were calculated for each scenario relative to the perceptual labels.

To compare discriminative performance, DeLong’s test for correlated ROC curves was applied within each outcome, accounting for the dependence between predictors derived from the same participants. Corresponding p values and 95% confidence intervals for AUC differences are reported in Table V.

**TABLE I:** ANOVA results from linear regression models for GoP at utterance and sound-class levels. SS = sum of squares; MS = mean squares; Resid. = residual; utt. = utterance.

| Model | Term | SS | MS | F | p |
| --- | --- | --- | --- | --- | --- |
| GoP (utt. level) | SD | 39.24 | 9.81 | 17.03 | < .001 |
|  | Gender | 0.07 | 0.07 | 0.12 | .726 |
|  | Age | 0.28 | 0.28 | 0.49 | .487 |
|  | Resid. | 85.84 | 0.58 |  |  |
| GoP (utt. level) | INT | 29.76 | 14.88 | 23.66 | < .001 |
|  | Gender | 0.08 | 0.08 | 0.13 | .721 |
|  | Age | 0.61 | 0.61 | 0.97 | .326 |
|  | Resid. | 94.97 | 0.63 |  |  |
| GoP (sc level) | SD | 77.38 | 19.35 | 31.95 | < .001 |
|  | SC | 8.46 | 8.46 | 13.97 | < .001 |
|  | Gender | 0.19 | 0.19 | 0.31 | .576 |
|  | Age | 0.59 | 0.59 | 0.98 | .324 |
|  | SD×SC | 0.44 | 0.11 | 0.18 | .949 |
|  | Resid. | 181.62 | 0.61 |  |  |
| GoP (sc level) | INT | 60.71 | 30.35 | 46.58 | < .001 |
|  | SC | 8.46 | 8.46 | 12.98 | < .001 |
|  | Gender | 0.11 | 0.11 | 0.16 | .686 |
|  | Age | 1.16 | 1.16 | 1.78 | .183 |
|  | INT×SC | 0.15 | 0.07 | 0.11 | .894 |
|  | Resid. | 198.10 | 0.65 |  |  |

**TABLE II:** ANOVA results from linear regression models for PostProb at utterance and sound-class levels.

| Model | Term | *SS* | *MS* | *F* | *p* |
| --- | --- | --- | --- | --- | --- |
| PP (utt. level) | SD | 5.38 | 1.35 | 11.46 | < .001 |
|  | Gender | 0.72 | 0.72 | 6.09 | .015 |
|  | Age | 0.30 | 0.30 | 2.59 | .110 |
|  | Resid. | 17.51 | 0.12 |  |  |
| PP (utt. level) | INT | 3.59 | 1.80 | 14.08 | < .001 |
|  | Gender | 1.05 | 1.05 | 8.20 | .005 |
|  | Age | 0.01 | 0.01 | 0.10 | .747 |
|  | Resid. | 19.26 | 0.13 |  |  |
| PP (sc level) | SD | 12.20 | 3.05 | 17.39 | < .001 |
|  | SC | 4.18 | 4.18 | 23.81 | < .001 |
|  | Gender | 1.43 | 1.43 | 8.16 | .005 |
|  | Age | 0.52 | 0.51 | 2.93 | .088 |
|  | SD×SC | 1.39 | 0.35 | 1.98 | .097 |
|  | Resid. | 52.63 | 0.18 |  |  |
| PP (sc level) | INT | 8.32 | 4.16 | 22.37 | < .001 |
|  | SC | 4.18 | 4.18 | 22.46 | < .001 |
|  | Gender | 2.14 | 2.14 | 11.48 | .001 |
|  | Age | 0.00 | 0.00 | 0.02 | .895 |
|  | INT×SC | 1.15 | 0.58 | 3.10 | .046 |
|  | Resid. | 56.56 | 0.19 |  |  |

**TABLE II:**
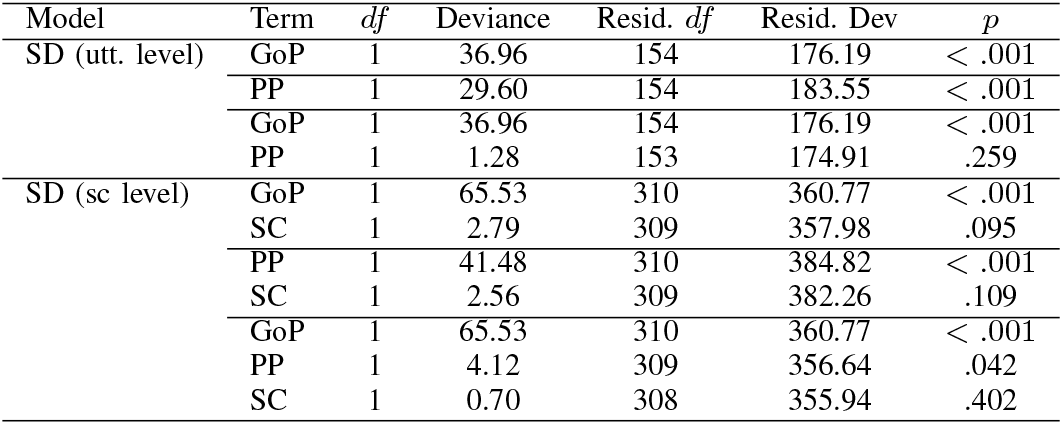
Logistic regression results for sound distortion at utterance and sound-class levels. df = degrees of freedom; Dev = deviance.

**TABLE IV:** Logistic regression results for intelligibility at utterance and sound-class levels.

| Model | Term | *df* | Deviance | Resid. *df* | Resid. Dev | *p* |
| --- | --- | --- | --- | --- | --- | --- |
| INT (utt. level) | GoP | 1 | 21.35 | 154 | 86.09 | < .001 |
|  | PP | 1 | 11.67 | 154 | 95.77 | < .001 |
|  | GoP | 1 | 21.35 | 154 | 86.09 | < .001 |
|  | PP | 1 | 0.41 | 153 | 85.68 | .521 |
| INT (sc level) | GoP | 1 | 41.14 | 310 | 173.75 | < .001 |
|  | SC | 1 | 0.87 | 309 | 172.88 | .351 |
|  | PP | 1 | 18.59 | 310 | 196.30 | < .001 |
|  | SC | 1 | 1.66 | 309 | 194.63 | .197 |
|  | GoP | 1 | 41.14 | 310 | 173.75 | < .001 |
|  | PP | 1 | 0.02 | 309 | 173.73 | .898 |
|  | SC | 1 | 1.01 | 308 | 172.72 | .316 |

**TABLE V:** ROC analysis results across speech units. Sens. = sensitivity; Spec. = specificity.

| Level | Metric | Rate | *AUC* | *SE* | *p* | Threshold | Sens. | Spec. |
| --- | --- | --- | --- | --- | --- | --- | --- | --- |
| Utterance | GoP | SD | 0.803 | 0.015 | < .001 | −0.391 | 0.753 | 0.718 |
|  |  | INT | 0.843 | 0.023 | < .001 | −0.763 | 0.941 | 0.688 |
|  | PP | SD | 0.778 | 0.016 | < .001 | −0.113 | 0.764 | 0.675 |
|  |  | INT | 0.781 | 0.029 | < .001 | −0.219 | 0.824 | 0.661 |
| Consonants | GoP | SD | 0.811 | 0.015 | < .001 | −0.380 | 0.865 | 0.632 |
|  |  | INT | 0.829 | 0.026 | < .001 | −0.973 | 0.824 | 0.730 |
|  | PP | SD | 0.745 | 0.017 | < .001 | −0.017 | 0.742 | 0.658 |
|  |  | INT | 0.719 | 0.033 | .001 | −0.221 | 0.706 | 0.709 |
| Vowels | GoP | SD | 0.777 | 0.016 | < .001 | 0.065 | 0.843 | 0.590 |
|  |  | INT | 0.848 | 0.021 | < .001 | −0.645 | 0.882 | 0.730 |
|  | PP | SD | 0.738 | 0.017 | < .001 | −0.277 | 0.685 | 0.701 |
|  |  | INT | 0.809 | 0.024 | < .001 | −0.431 | 0.824 | 0.672 |

### F. Statistical Analysis

Mean and standard deviation values from all control participants were used to z-normalize GoP scores and log-odds posterior probabilities (PP). Pairwise comparisons were conducted using Wilcoxon tests, with p values adjusted for multiple comparisons using the Benjamini–Hochberg procedure for both sound distortion (SD) and intelligibility (INT). Within the MSD group, separate linear regression models were fitted with z-transformed GoP scores and log-odds posterior probabilities as outcomes and sound distortion or intelligibility as predictors, with gender and age included as covariates. Analyses were performed at both the utterance and sound class levels to assess global and segment-specific patterns. At the utterance level, the model specification was as follows:

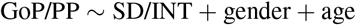

At the sound-class level, interaction terms between sound distortion or intelligibility and sound class (SC, consonants vs vowels) were included to assess whether associations differed across sound classes. The model was specified as follows:

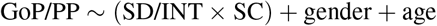

All categorical predictors were dummy coded using treatment contrasts, with “normal” as the reference level for both sound distortion and intelligibility. Post hoc pairwise comparisons across levels of sound distortion and intelligibility were conducted with p values adjusted using the same procedures applied in the Wilcoxon tests.

Using the same dataset as in the ROC analysis, ordinal logistic regression models were fit to evaluate the extent to which metrics predict perceptual severity at the utterance level and at the sound-class level. For these analyses, both perceptual ratings were recoded as binary variables, with sound distortion categorized as normal (normal and equivocal) versus unequivocally abnormal (mild, moderate, marked, and severe), and intelligibility categorized as normal (normal and equivocally reduced) versus unequivocally abnormal. Separate models were specified with binarized sound distortion or intelligibility as outcomes, predicted by z-transformed GoP, z-transformed log-odds posterior probabilities, or both (see the formula below). For sound-class-level analyses, sound class was included as an additional predictor. The model specification was as follows:

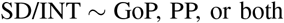

All analyses were conducted in R [31]. Linear mixed-effects models were fit using the `lme4` package [32], with significance testing performed using `lmerTest` [33]. Estimated marginal means and post hoc comparisons were obtained using `emmeans` [34]. Logistic regression models were fit using the `glm()` function in the base R `stats` package.

## III. Results

### A. GoP

As shown in Fig. 1, transformer-based representations consistently outperform traditional acoustic baselines for both sound distortion and intelligibility. For sound distortion (Fig. 1A), the highest Kendall’s *τ* values obtained using Mel-spectrogram and MFCC features are 0.399 and 0.404, respectively. WavLM features achieve substantially higher correlations across all classifiers, with the highest mean correlations obtained for GM (0.491) and kNN (0.489). The highest overall correlation is 0.518, achieved by kNN at layer 23, while PSVM and GM reach comparable peak values of 0.515 and 0.511, respectively. XLS-R also outperforms the acoustic baselines, with the highest mean correlations of 0.484 (GM) and 0.469 (kNN), and a maximum value of 0.510 obtained by GM at layer 19.

**Fig. 1:**
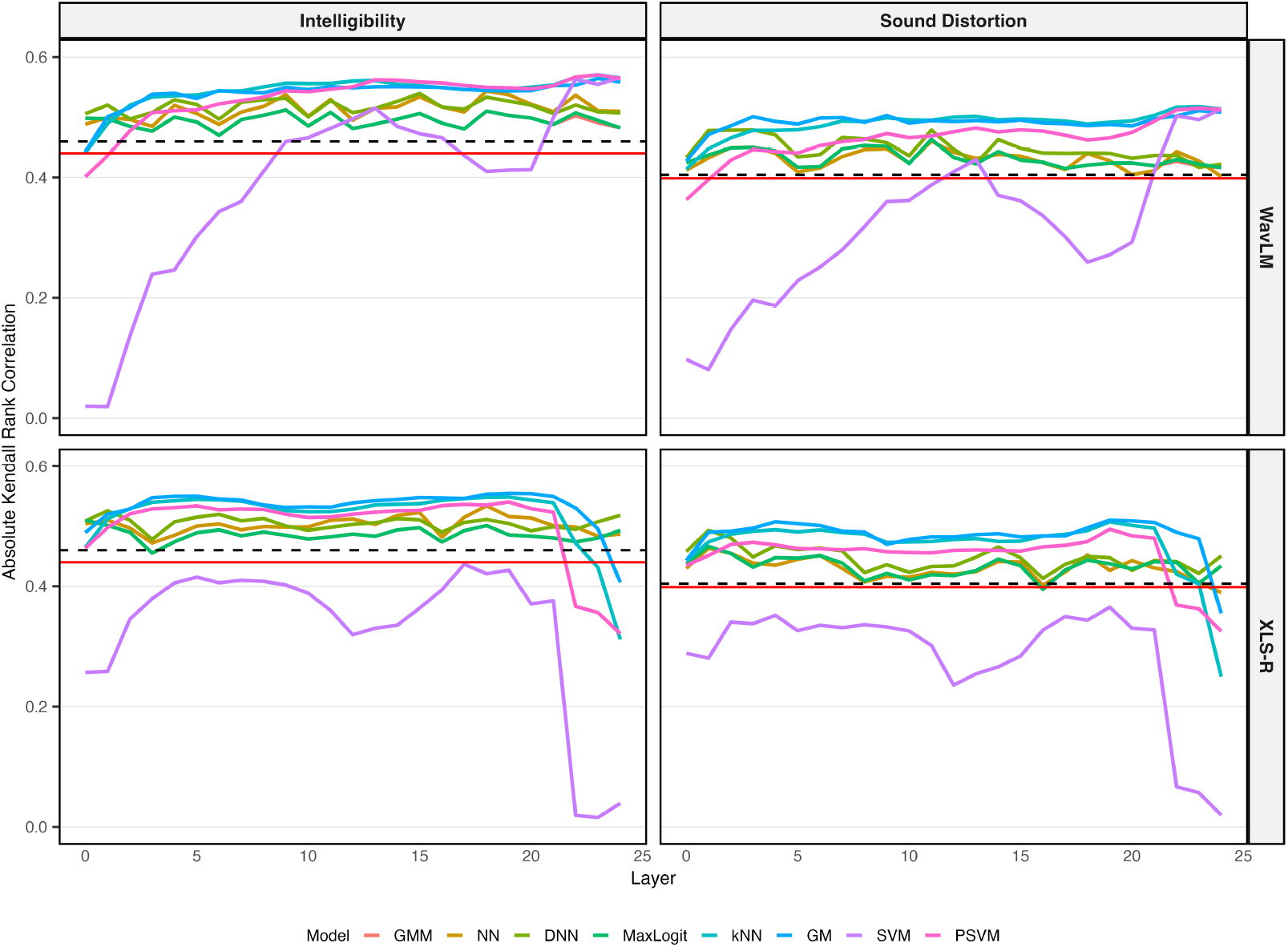
Absolute Kendall’s *τ* correlations between GoP scores and perceptual ratings for features extracted across layers of S3M models. The dashed black line indicates the maximum correlation for Mel-spectrogram features, and the solid red line indicates the maximum correlation for MFCC features.

For intelligibility (Fig. 1), the highest correlations obtained with Mel-spectrogram and MFCC features are 0.440 and 0.460, respectively. WavLM again provides the strongest performance, with the highest mean correlations achieved by kNN (0.543), GM (0.540), and PSVM (0.532). The highest overall correlation is 0.570, obtained by PSVM at layer 23, while kNN achieves a nearly identical value (0.570) at the same layer. XLS-R follows a similar trend, with GM (0.532) and kNN (0.518) yielding the highest mean correlations, and a maximum value of 0.554 achieved by GM at layer 19. Across both transformer models, SVM consistently exhibits the lowest mean correlations despite achieving competitive peak values at individual layers.

Based on these results, kNN is selected as the final model because it provides the strongest overall performance across both perceptual ratings. It achieves the highest correlation for sound distortion (*τ* = 0.518) and a nearly identical highest correlation for intelligibility (*τ* = 0.570). To ensure a unified feature representation for both prediction tasks, we select WavLM layer 23, as it is the layer at which kNN achieves its highest correlations for both sound distortion and intelligibility.

At the utterance level (Fig. 2A–B), pairwise comparisons indicated that, for sound distortion, GoP mean in the controls did not differ from the normal condition in the MSD group but was significantly higher than all other distortion levels (*p*s ¡ .05). Correspondingly, GoP standard deviation showed significant differences for the mild and moderate conditions (*ps* < .01). For intelligibility, GoP mean in the controls was significantly higher than all MSD levels (all *ps* < .001), with significant differences in standard deviation observed for the equivocal and abnormal conditions (*ps* < .01). At this level, gender effects were minimal (Fig. 2C), and age showed weak associations with substantial variability (Fig. 2D).

**Fig. 2:**
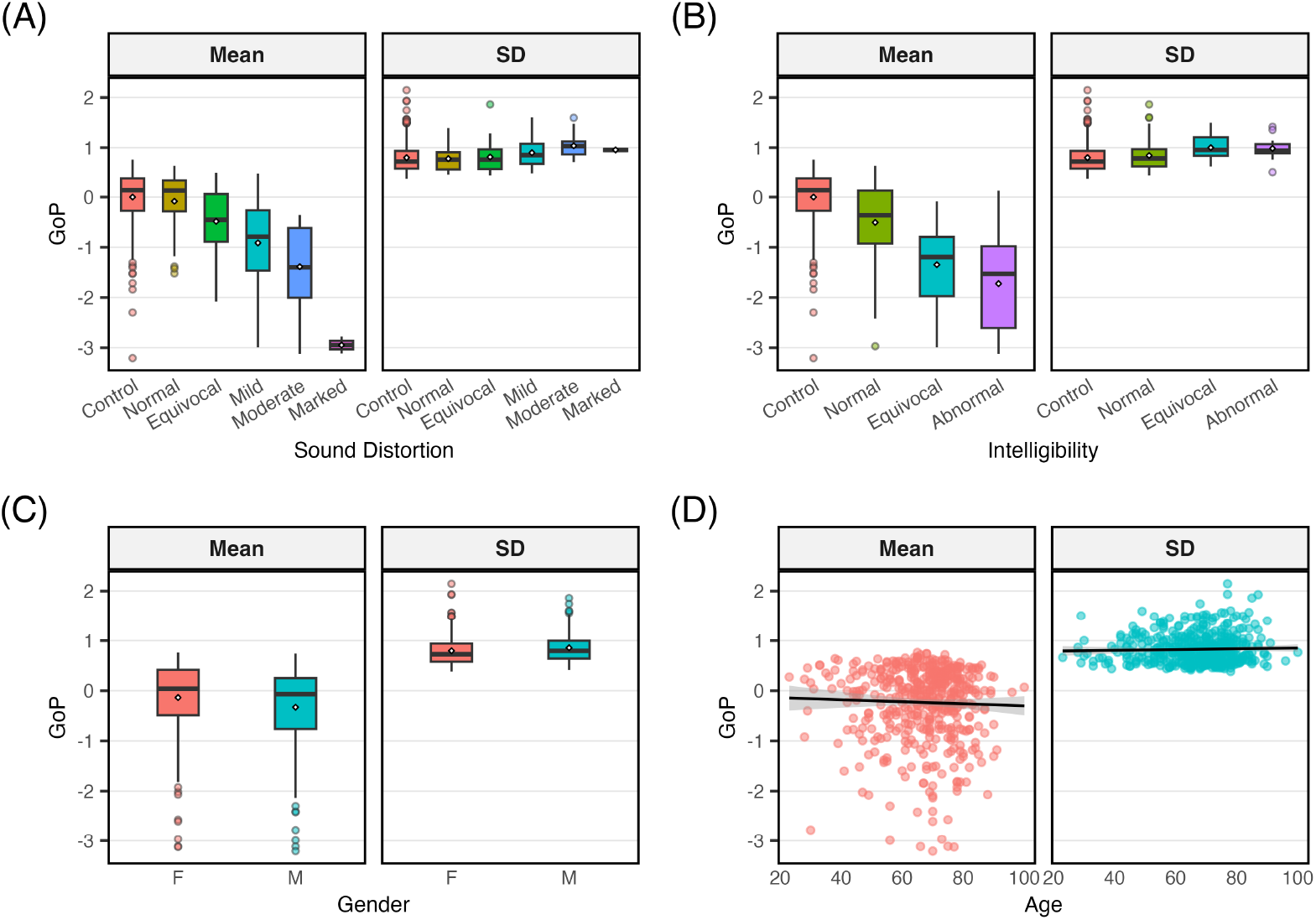
Distributions of z–normalized, utterance-level kNN-derived GoP mean and standard deviation by sound distortion (A), intelligibility (B), gender (C), and age (D); means are indicated by diamonds (A–C) and standard error by grey shading (D).

Linear model analyses conducted at the utterance level confirmed a significant effect of sound distortion on GoP scores (*F* (4, 149) = 17.03, *p* < .001), and intelligibility (*F* (2, 151) = 23.66, *p* < .001; see Table I). Follow-up pairwise comparisons indicated a graded decline in GoP scores with increasing severity. For sound distortion, all contrasts were significant (all *ps* < .05). For intelligibility, all contrasts were significant (*ps* < .001), except for equivocal vs. abnormal.

At the sound class level (Fig. 3A–B), GoP mean decreased with increasing sound distortion and intelligibility severity for both consonants and vowels (*ps* < .05), except for control vs. normal for sound distortion. For GoP standard deviation, fewer differences were observed: for sound distortion, control only differed from mild and moderate conditions for both consonants and vowels; for intelligibility, control differed from equivocal and abnormal for both vowels and consonants. Consistent with the patterns observed at the utterance level, gender effects remained minimal and age still showed weak, variable trends (Fig. 3C–D).

**Fig. 3:**
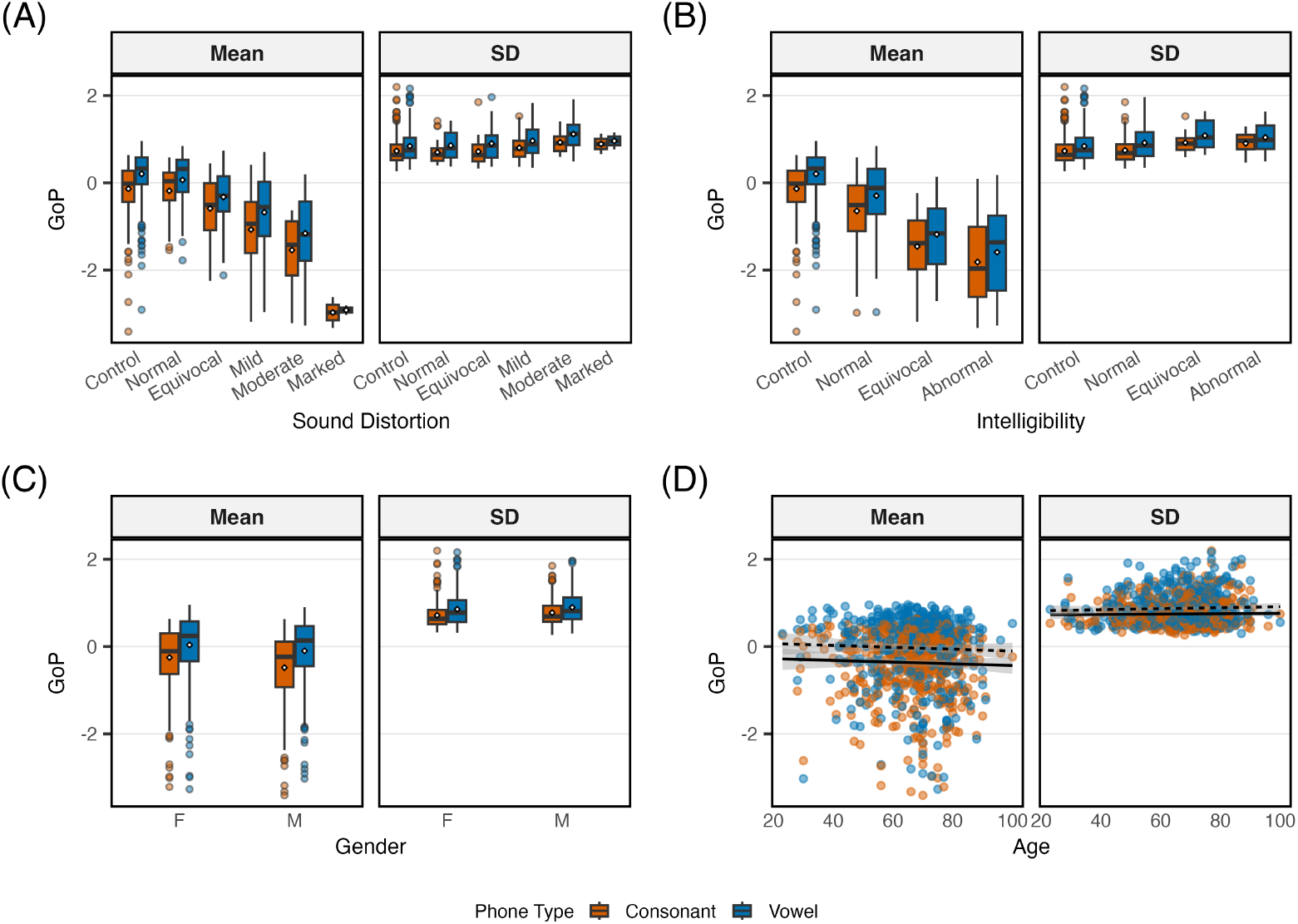
Distributions of z–normalized, sound-class-level kNN-derived GoP mean and standard deviation by sound distortion (A), intelligibility (B), gender (C), and age (D); means are indicated by diamonds (A–C) and standard error by grey shading (D).

Linear model analyses conducted at the sound class level confirmed a significant effect of sound distortion (*F* (4, 300) = 31.95, *p* < .001), and intelligibility (*F* (2, 304) = 46.58, *p* < .001; see Table I), as well as a significant main effect of phone type (*ps* < .001), while the interaction between severity and phone type was not significant. Follow-up pairwise comparisons indicated a graded decline across severity levels, with all contrasts significant for sound distortion (all *ps* < .01) and all contrasts significant for intelligibility (*ps* ≤ .001) except equivocal vs. abnormal. Gender and age were not significant predictors for both perceptual ratings.

### B. Phonet

Similar to GoP, posterior probabilities showed stronger associations with sound distortion (0.347) than with intelligibility (0.287). At the utterance level (Fig. 4A-B), mean of posterior probabilities showed a systematic reduction with increasing sound distortion and intelligibility severity. For sound distortion, values in the control group were comparable to the normal condition but exceeded all remaining conditions (*ps* < .05). For intelligibility, controls displayed higher values than all conditions (all *ps* < .001), indicating clear separa-tion. In contrast, standard deviation of posterior probabilities exhibited no differentiation, with all comparisons not reaching significance, except for control vs. abnormal for intelligibility. With respect to speaker characteristics, gender effects were present but small (Fig. 4C), whereas age showed weak and variable trends (Fig. 4D).

**Fig. 4:**
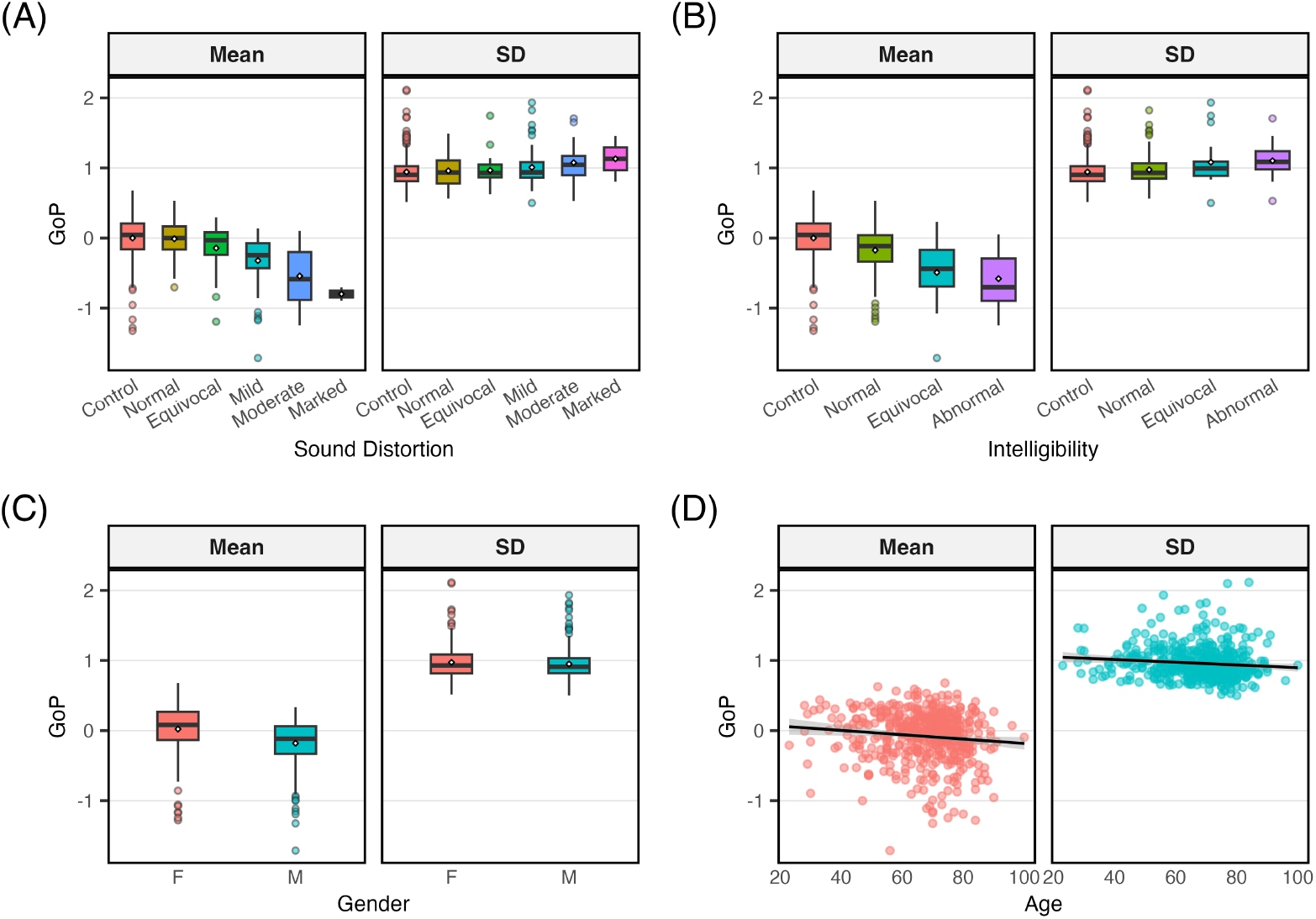
Distributions of utterance-level z–normalized log-odds posterior probabilities (PostProb) mean and standard deviation by sound distortion (A), intelligibility (B), gender (C), and age (D); means are indicated by diamonds (A–C) and standard error by grey shading (D).

Linear model analyses conducted at the utterance level revealed significant effects of sound distortion on phonological posterior scores (*F* (4, 149) = 11.46, *p* < .001), as well as intelligibility (*F* (2, 151) = 14.08, *p* < .001; see Table II). Gender was also a significant predictor in both models, including those with sound distortion (*F* (1, 149) = 6.09, *p* < .05), and intelligibility (*F* (1, 151) = 8.20, *p* < .01), with males exhibiting lower posterior scores than females. Follow-up pairwise comparisons demonstrated a graded reduction in posterior scores with increasing severity. For sound distortion, most contrasts reached significance (*ps* < .05), except for normal vs. equivocal and mild/moderate vs. marked. For intelligibility, all contrasts were significant (*ps* ≤ .01), except for equivocal vs. abnormal.

At the sound class level (Fig. 5A–B), mean of posterior probabilities similarly declined as severity increased, while also revealing differences between consonants and vowels. For sound distortion, control was significantly higher than mild and moderate conditions for consonants (*ps* < .001), while for vowels, significant differences were observed for all conditions (*ps* < .05), except for normal. For intelligibility, control was significantly higher than all conditions for both consonants and vowels (all *ps* < .001). Patterns for standard deviation of posterior probabilities were less pronounced. No significant differences were observed for sound distortion across either sound class. For intelligibility, standard deviation differences were largely non-significant, except for control vs. abnormal for vowels (*p* < .01). This contrast between vowels and consonants remained evident across gender and age, with minimal gender-related variation and weak age-related effects (Fig. 5C–D).

**Fig. 5:**
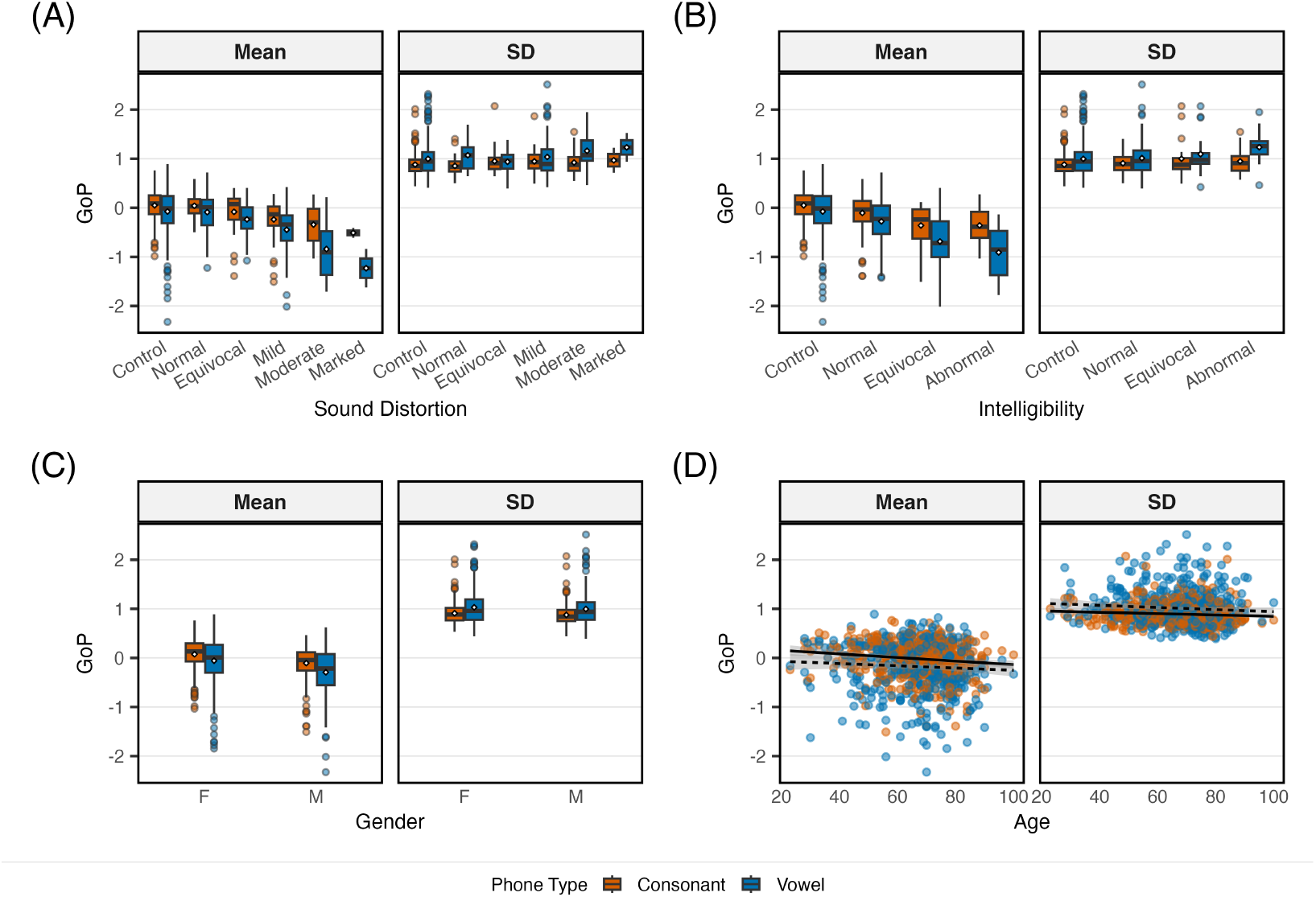
Distributions of sound-class-level z–normalized log-odds posterior probabilities (PostProb) mean and standard deviation by sound distortion (A), intelligibility (B), gender (C), and age (D); means are indicated by diamonds (A–C) and standard error by grey shading (D).

Linear model analyses conducted at the sound class level further supported these patterns, showing significant effects of sound distortion (*F* (4, 300) = 17.39, *p* < .001), and intelligibility (*F* (2, 304) = 22.37, *p* < .001; see Table II). In addition, sound class (*ps* < .001) and gender (*ps* < .01 were significant predictors, while age did not emerge as a significant predictor. The interaction between sound distortion and sound class was not significant, whereas a significant interaction between intelligibility and sound class was observed (*F* (2, 304) = 3.10, *p* < .05), indicating that intelligibility effects differed between consonants and vowels. Follow-up comparisons showed a stepwise separation across severity conditions for sound distortion, with most contrasts reaching significance, except for normal vs. equivocal and moderate vs. marked. For intelligibility, significant differences were primarily observed for vowels, including normal vs. equivocal and normal vs. abnormal (*ps* < .01), whereas no contrasts reached significance for consonants.

### C. GoP vs. Phonet

Across both utterance- and sound-class-level analyses, logistic regression models showed that GoP scores and posterior probabilities significantly predicted the severity of sound distortion and intelligibility (all *ps* < .001; see Tables III and IV),with higher values associated with a lower likelihood of belonging to the abnormal condition. At both levels, each predictor was significant when considered separately. However, when included jointly, GoP scores remained a significant predictor across conditions (utterance-level: *β* = −1.06, *SE* = 0.38, *z* = −2.84, *p* < .01; sound-class-level: *β* = −1.07, *SE* = 0.22, *z* = −4.80, *p* < .001 for sound distortion; utterance-level: *β* = −1.58, *SE* = 0.54, *z* = −2.91, *p* < .01; sound-class-level: *β* = − 1.35, *SE* = 0.30, *z* = − 4.46, *p* < .001 for intelligibility), whereas posterior probabilities did not provide additional explanatory power. A small improvement in model fit was observed only for sound distortion at the sound-class level (*χ*^2^(1) = 4.12, *p* < .05), but not for intelligibility.

ROC analyses (Fig. 6) showed that GoP consistently out-performed posterior probabilities in discriminating both sound distortion and intelligibility across speech units (see Table V). At the utterance level, GoP achieved higher AUCs than posterior probabilities for sound distortion (0.803 vs. 0.778) and intelligibility (0.843 vs. 0.781). This advantage was preserved at the sound-class level. For consonants, AUCs were 0.811 for GoP and 0.745 for posterior probabilities for sound distortion, and 0.829 versus 0.719 for intelligibility. For vowels, AUCs were 0.777 versus 0.738 for sound distortion and 0.848 versus 0.809 for intelligibility. Overall, classification performance was consistently higher for intelligibility than for sound distortion, and GoP provided more accurate discrimination than posterior probabilities across all speech units.

**Fig. 6:**
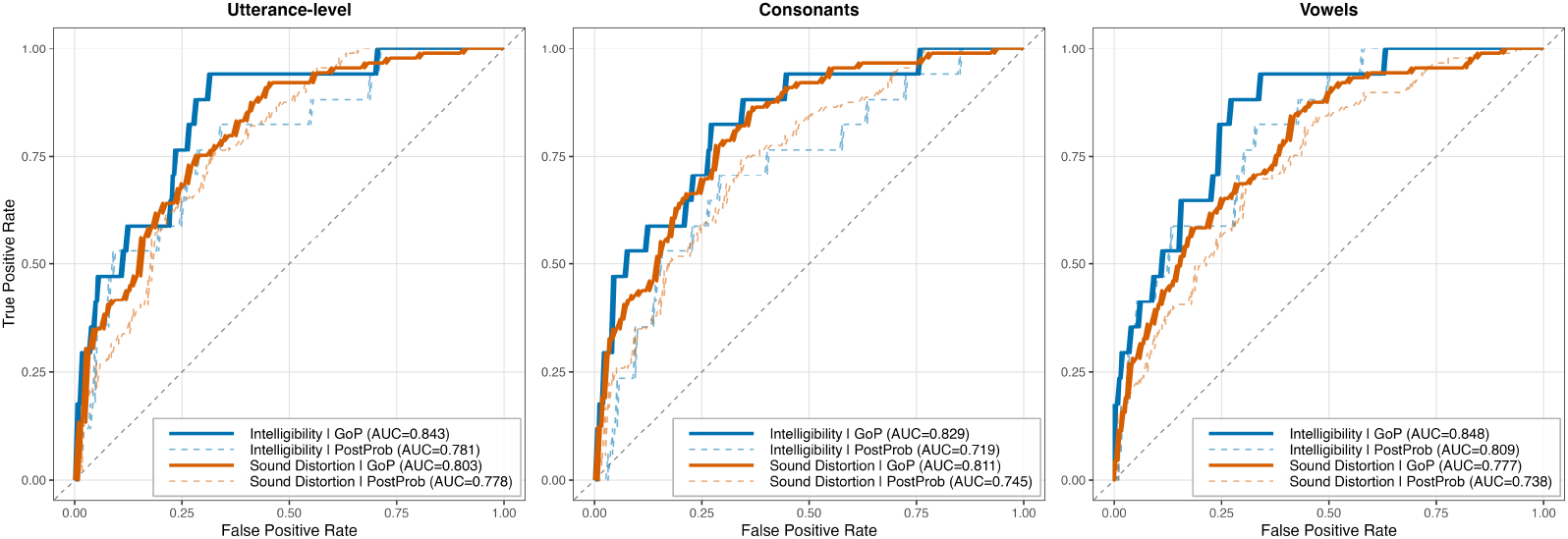
ROC curves of GoP and posterior probability for intelligibility and sound distortion across speech units.

Sensitivity–specificity trade-off analyses (Fig. 7) revealed consistent patterns across speech units and perceptual ratings. The OR condition yielded the highest sensitivity across all cases. At the utterance level, sensitivity reached 0.865 for sound distortion and 0.941 for intelligibility, compared with 0.753 and 0.941 for GoP alone and 0.764 and 0.824 for posterior probabilities alone, respectively. In contrast, the AND condition achieved the highest specificity, reaching 0.812 for sound distortion and 0.772 for intelligibility at the utterance level, but at the expense of lower sensitivity (0.652 and 0.824, respectively). Similar trade-offs were observed for consonants and vowels. For sound distortion, OR increased sensitivity to 0.910 for both consonants and vowels, whereas AND increased specificity to 0.812 and 0.803, respectively. For intelligibility, OR achieved sensitivities of 0.882 for both consonants and vowels, while AND yielded the highest specificities (0.820 and 0.810, respectively). Overall, GoP alone provided the most balanced sensitivity–specificity profile, consistently outperforming posterior probabilities while avoiding the pronounced trade-off observed with the OR and AND fusion strategies.

**Fig. 7:**
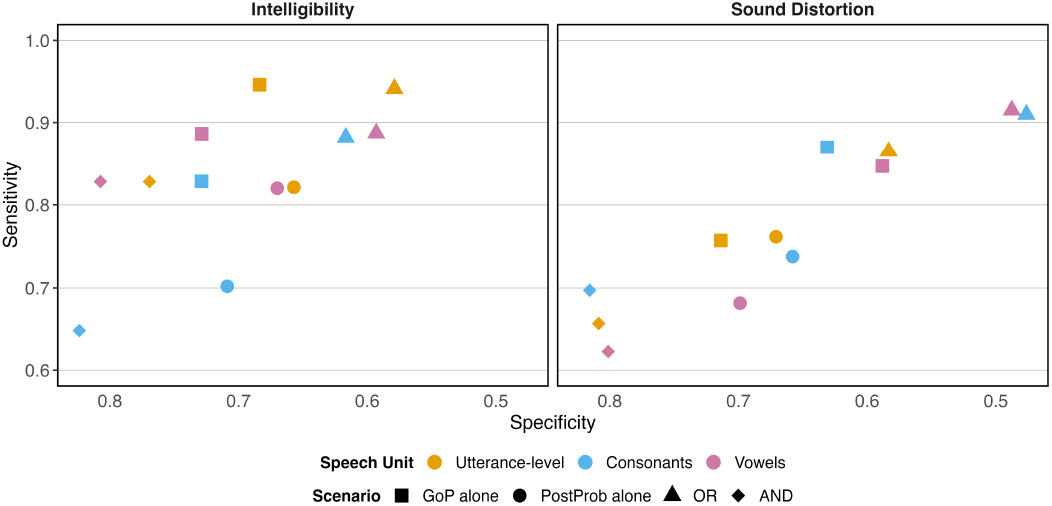
Sensitivity–specificity trade-off for intelligibility and sound distortion across speech units and decision scenarios.

As illustrated in Fig. 8, GoP and posterior probabilities exhibited a clear positive relationship for both sound distortion and intelligibility across all speech units. Higher GoP values were generally associated with higher posterior probabilities, indicating that the two measures provide complementary information for distinguishing perceptual outcomes. For both perceptual ratings, normal (or absent) tokens were predominantly clustered in the upper-right region of the plots, whereas abnormal (or present) tokens were concentrated toward the lower-left, although some overlap was observed near the decision boundaries. This overall pattern was consistent at the utterance, consonant, and vowel levels. The dashed lines indicate the ROC-derived thresholds for GoP and posterior probabilities, illustrating how combining the two measures can improve discrimination by separating most correctly classified tokens into the expected quadrants while highlighting ambiguous cases near the thresholds.

**Fig. 8:**
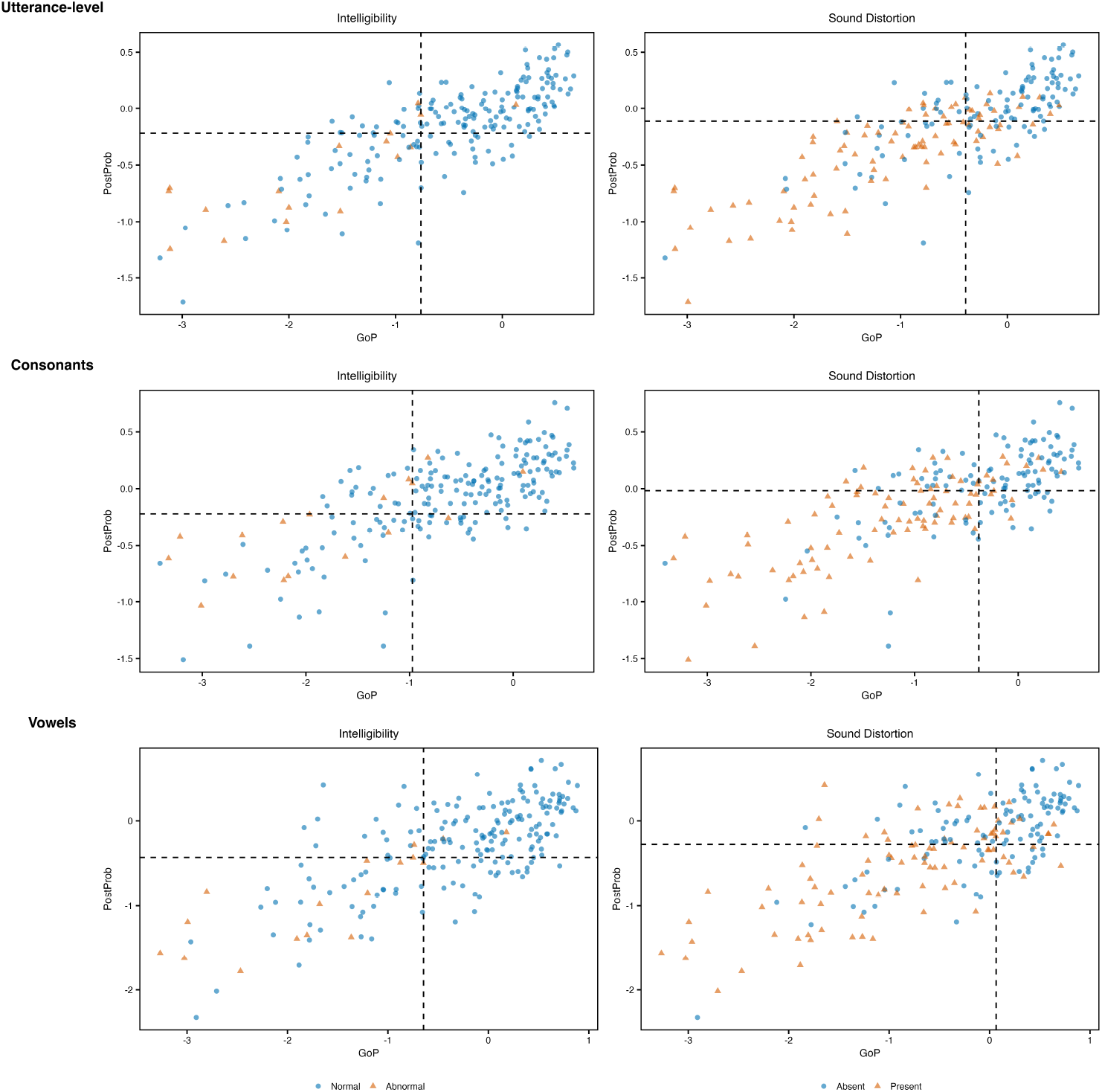
Relationship between GoP and posterior probability for binarized intelligibility and sound distortion across speech units, with dashed lines indicating ROC-derived classification thresholds.

To further examine disagreements between GoP- and posterior probability-based predictions, phone-level trajectories were grouped into mismatch categories based on whether each measure agreed with the perceptual labels for intelligibility (INT) and sound distortion (SD). No cases were observed in which both measures incorrectly predicted a normal/absent outcome for an abnormal/present token. Figure 9 therefore presents the remaining mismatch categories, including cases where both measures incorrectly predicted abnormal/present and cases where only GoP or posterior probabilities disagreed with the perceptual labels.

**Fig. 9:**
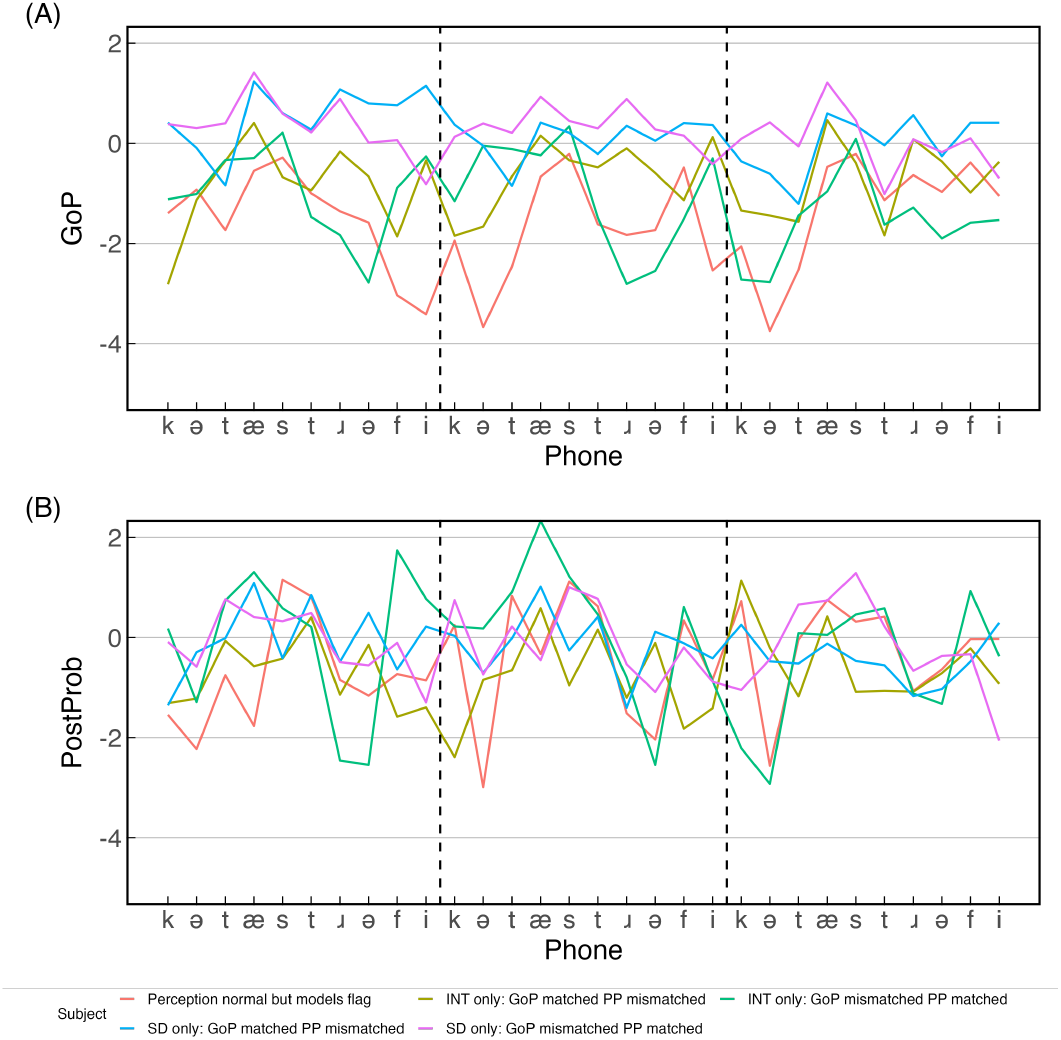
Phone-level changes in GoP (A) and posterior probabilities (B) across three repetitions of the word catastrophe for selected participants, shown by mismatch category at the utterance level. Dashed vertical lines indicate boundaries between repetitions.

Figure 9 shows phone-level trajectories of GoP (A) and posterior probabilities (B) across three repetitions of catastrophe for the observed mismatch categories. Overall, GoP exhibited clearer separation between mismatch categories than posterior probabilities. Consonants showed relatively stable trajectories with modest differences across categories, whereas vowels exhibited greater variability and clearer separation, particularly for the central vowels. In contrast, posterior probability trajectories displayed greater overlap across categories for both consonants and vowels, with no consistent separation across phones. These findings are consistent with the ROC analyses, indicating that GoP provides more discriminative information than posterior probabilities for both sound distortion and intelligibility.

## IV. Discussion

This study examined the objective characterization of segmental speech deviation in speakers with and without motor speech disorders, focusing on how GoP and phonological posterior probabilities relate to perceptual ratings of sound distortion and intelligibility. We also evaluated the influence of age and gender on these relationships. By systematically varying feature representations and GoP formulations, and directly comparing phoneme-based and articulatory feature–based approaches, we show that both measures are sensitive to clinically relevant variation in speech production, although GoP consistently demonstrated stronger associations with perceptual ratings than phonological posterior probabilities, indicating that the two measures capture overlapping but complementary aspects of speech impairment.

### A. Impact of Self-Supervised Representations on GoP

The first research question examined whether S3M representations improve GoP-based assessment relative to traditional acoustic features. The present findings provide clear evidence that S3M representations, particularly WavLM, substantially strengthen the relationship between GoP scores and perceptual ratings of both sound distortion and intelligibility [12], [15], [35]. This improvement suggests that S3M representations capture speech variability more effectively than conventional acoustic features. Traditional acoustic features, such as MFCCs and Mel spectrograms, primarily capture short-term spectral properties and may therefore be limited in their ability to represent the context-dependent and gradient nature of articulatory deviations in MSD [14], [23]. In contrast, S3M representations encode hierarchical information that integrates local acoustic detail with broader temporal and contextual structure, allowing for a more robust characterization of coarticulation and subtle distortions [15], [16].

The observed advantage of higher-layer representations further indicates that contextualized embeddings are particularly informative for pronunciation assessment in clinical populations. These findings are consistent with the idea that higher-layer representations encode more abstract phonetic and phonological information [12], [18], enabling improved discrimination between typical variation and clinically meaningful deviation. In addition, the strong performance of kNN-based GoP suggests that the strong performance of kNN suggests that nonparametric approaches may be better suited to modeling atypical speech, as they do not rely on strict assumptions about phoneme distributions and can accommodate the increased variability and overlap characteristic of MSD [13], [25].

Taken together, these findings highlight that effective GoP-based assessment depends on both rich representation learning and flexible modeling strategies. They demonstrate the value of integrating S3M features with approaches that can accommodate noncanonical speech patterns.

### B. Relationship Between GoP and Phonological Posterior Features

The second research question investigated how GoP and phonological posterior probabilities relate to perceptual ratings of speech impairment, and whether they reflect overlapping or distinct indices of severity. The results indicate that both measures are significantly associated with sound distortion and intelligibility, but differ in the type and strength of information they capture and how they relate to perceptual judgments [36]. Consistent with earlier results, GoP derived from WavLM representations, particularly from higher model layers using kNN, showed the most robust performance across perceptual ratings.

GoP operates within a phoneme classification framework, quantifying the degree of deviation from expected phoneme realizations. When entered into the same severity classification models, GoP consistently outperformed phonological posterior probabilities across the correlation, regression, and ROC analyses. This dominance likely reflects the intrinsic relationship between perceptual intelligibility and phoneme-level categorization: intelligibility judgments are fundamentally tied to whether phonemic contrasts are preserved sufficiently for lexical access [37]. Importantly, while GoP aligns closely with intelligibility judgments, intelligibility itself is never used in model training. GoP models are trained exclusively on acoustic–phonetic representations derived from typical speech. The observed associations therefore likely reflect shared sensitivity to phoneme-level disruption rather than direct optimization for the predicted perceptual labels. In contrast, phonological posterior probabilities provide a feature-based representation of speech that reflects articulatory configuration rather than categorical phoneme identity, offering a more direct account of how speech is produced [38]; these may provide complementary insight into articulatory configuration, particularly in cases where phoneme-level measures alone may not fully capture underlying motor speech differences.

This distinction is further supported by the mismatch analyses, which suggest that GoP and phonological posterior probabilities capture partially distinct aspects of speech impairment. Cases in which GoP detects abnormality while phonological posterior probabilities do not may indicate deviations that disrupt phoneme-level categorization without substantially altering articulatory feature patterns. Conversely, cases in which phonological posterior probabilities detect abnormality while GoP does not may reflect more subtle articulatory differences that preserve phoneme identity despite underlying motor impairment.

### C. Effects of Age and Gender

The third research question examined whether age and gender influence model-derived measures or their association with perceptual severity. The findings suggest that these factors have minimal impact on the relationship between model outputs and perceptual ratings, despite their known effects on acoustic characteristics of speech [21], [22].

Age and gender are well established to influence low-level acoustic properties, including fundamental frequency, formant structure, and spectral characteristics [39], [40]. However, the relative stability of both GoP and phonological posterior measures across these factors indicates that they capture aspects of speech that are robust to speaker-specific variability [21], [22]. This robustness may partly reflect the use of S3M representations, which are trained on large and diverse datasets and are therefore better able to abstract away from demographic variation and emphasize linguistically and clinically relevant information [6], [15].

Although a small but statistically significant gender effect was observed for phonological posterior probabilities, this effect did not meaningfully alter their relationship with perceptual severity. From a clinical perspective, this relative invariance is encouraging, as it supports the potential applicability of these measures across diverse populations without requiring extensive demographic normalization.

### D. Limitations and Future Directions

Several limitations should be considered. First, speech samples were limited to repeated productions of a single word, which may restrict generalizability to more naturalistic speech contexts. However, the use of a fixed utterance may also offer practical advantages in clinical settings, particularly for individuals with more severe motor speech or cognitive impairments, by reducing task demands and enabling more consistent comparisons across recordings [1], [7]. Future work should evaluate both approaches using connected speech to determine whether they provide complementary or differentially specific information for assessing speech impairment [6]. Second, although a range of severity levels was included, fewer observations were available at more severe levels, which limited our ability to assess floor effects of GOP and phono-logical posteriors. Third, because both GoP and phonological posterior measures rely on forced alignment, segmentation errors may influence both segment-level and aggregated scores. Misalignment could introduce systematic bias, particularly for shorter or acoustically variable segments. Future work should assess sensitivity to alignment quality. Fourth, while this study focused on GoP and phonological posterior probabilities, additional approaches, including end-to-end models and multimodal representations, may further inform understanding of speech impairment. Fifth, demographic effects were examined in a limited scope, and future studies should consider additional sources of variability such as dialect, language background, and disease subtype [21]. Finally, the present study was not powered for subtype-specific analyses. Different types of motor speech disorders may differentially affect GoP and phonological posterior measures, reflecting variation in phoneme-level disruption and articulatory impairment. Future studies incorporating subtype-balanced cohorts, longitudinal data, and connected speech samples will be important for determining whether these measures are differentially sensitive to specific motor speech disorder profiles.

## IV. Conclusion

This study demonstrates that both GoP and phonological posterior probabilities provide meaningful and clinically relevant measures of speech impairment in individuals with MSD. Across modeling approaches, self-supervised speech representations, particularly WavLM, substantially improved the ability to capture variation in perceptual ratings of sound distortion and intelligibility, highlighting the importance of representation learning for objective speech assessment. GoP consistently outperformed phonological posterior probabilities across correlation, regression, and classification analyses, demonstrating stronger agreement with perceptual judgments of both sound distortion and intelligibility. In contrast, phonological posterior probabilities offered complementary insight into articulatory patterns of speech production but contributed limited additional predictive value when considered alongside GoP. These findings suggest that the two approaches capture partially overlapping but distinct aspects of speech impairment. Importantly, both approaches were relatively robust to age and gender, supporting their potential applicability across diverse clinical populations. Overall, these findings support the use of self-supervised GoP as an objective measure of speech impairment while highlighting the complementary role of phonological posterior probabilities in characterizing articulatory aspects of motor speech disorders.

## Data Availability

The datasets of the current study are available from the corresponding author on reasonable request.

## Acknowledgments

The authors thank Ashley D. Bachman for project administration and also thank the patients and caregivers who have generously committed themselves to this research.

## Footnotes

1 https://github.com/juice500ml/acoustic-units-for-ood

2 https://github.com/jcvasquezc/phonet

## References

[1] J. R. Duffy, Motor Speech Disorders: Substrates, Differential Diagnosis, and Management, 4th ed. St. Louis, MO, USA: Elsevier, 2020.

[2] M. Walshe, N. Miller, M. Leahy, and A. Murray, “Intelligibility of dysarthric speech: Perceptions of speakers and listeners,” Int. J. Lang. Commun. Disord., vol. 43, no. 6, 2008, doi: 10.1080/13682820801887117.

[3] K. J. Ballard, J. P. Granier, and D. A. Robin, “Understanding the nature of apraxia of speech: Theory, analysis, and treatment,” Aphasiology, vol. 14, no. 10, 2000, doi: 10.1080/02687030050156575.

[4] K. Bunton, R. D. Kent, J. R. Duffy, J. C. Rosenbek, and J. F. Kent, “Listener agreement for auditory-perceptual ratings of dysarthria,” J. Speech, Lang., Hear. Res., vol. 50, no. 6, 2007, doi: 10.1044/1092-4388(2007/102).

[5] A. Favaro et al., “Interpretable speech features vs. DNN embeddings: What to use in the automatic assessment of Parkinson’s disease in multi-lingual scenarios,” Comput. Biol. Med., vol. 166, 2023, doi: 10.1016/j.compbiomed.2023.107559.

[6] S.-I. Ng et al., “A tutorial on clinical speech AI development: From data collection to model validation,” 2024, doi: 10.48550/arXiv.2410.21640.

[7] M. Pernon, F. Assal, I. Kodrasi, and M. Laganaro, “Perceptual classification of motor speech disorders: The role of severity, speech task, and listener’s expertise,” J. Speech, Lang., Hear. Res., vol. 65, no. 8, 2022, doi: 10.1044/2022JSLHR-21-00519.

[8] S. Dudy, S. Bedrick, M. Asgari, and A. Kain, “Automatic analysis of pronunciations for children with speech sound disorders,” Comput. Speech Lang., vol. 50, 2018, doi: 10.1016/j.csl.2017.12.006.

[9] F. Wang, J. R. Duffy, A. D. Bachman, L. R. Barnard, H. Botha, and R. L. Utianski, “Deep learning-derived measures of sound-level accuracy in primary progressive apraxia of speech: A feasibility pipeline with descriptive evidence from two cases,” Clin. Linguist. Phon., 2026, doi: 10.1080/02699206.2026.2639397.

[10] S. M. Witt and S. J. Young, “Phone-level pronunciation scoring and assessment for interactive language learning,” Speech Commun., vol. 30, no. 2–3, 2000, doi: 10.1016/S0167-6393(99)00044-8.

[11] W. Hu, Y. Qian, F. K. Soong, and Y. Wang, “Improved mispronunciation detection with deep neural network trained acoustic models and transfer learning based logistic regression classifiers,” Speech Commun., vol. 67, 2015, doi: 10.1016/j.specom.2014.12.008.

[12] K. Choi, E. Yeo, K. Chang, S. Watanabe, and D. Mortensen, “Leveraging allophony in self-supervised speech models for atypical pronunciation assessment,” arXiv, 2025, doi: 10.48550/arXiv.2502.07029.

[13] E. Yeo, K. Choi, S. Kim, and M. Chung, “Speech intelligibility assessment of dysarthric speech by using goodness of pronunciation with uncertainty quantification,” arXiv, 2023, doi: 10.48550/arXiv.2305.18392.

[14] X. Xu, Y. Kang, S. Cao, B. Lin, and L. Ma, “Explore wav2vec 2.0 for mispronunciation detection,” in Proc. Interspeech 2021, 2021, pp. 4428–4432, doi: 10.21437/Interspeech.2021-777. [Online]. Available: https://www.isca-archive.org/interspeech2021/xu21kinterspeech.pdf

[15] S. Chen et al., “WavLM: Large-scale self-supervised pre-training for full stack speech processing,” 2021, doi: 10.48550/arXiv.2110.13900.

[16] A. Babu et al., “XLS-R: Self-supervised cross-lingual speech representation learning at scale,” arXiv, 2021, doi: 10.48550/arXiv.2111.09296.

[17] T.-H. Feng et al., “SUPERB @ SLT 2022: Challenge on generalization and efficiency of self-supervised speech representation learning,” 2022, doi: 10.48550/arXiv.2210.08634.

[18] X. Chang et al., “Exploring speech recognition, translation, and understanding with discrete speech units: A comparative study,” 2023, doi: 10.48550/arXiv.2309.15800.

[19] J. C. Vásquez-Correa, P. Klumpp, J. R. Orozco-Arroyave, and E. Nöth, “Phonet: A tool based on gated recurrent neural networks to extract phonological posteriors from speech,” in Proc. Interspeech 2019, 2019, pp. 549–553, doi: 10.21437/Interspeech.2019-1405.

[20] M. Cernak, J. R. Orozco-Arroyave, F. Rudzicz, H. Christensen, J. C. Vásquez-Correa, and E. Nöth, “Characterisation of voice quality of Parkinson’s disease using differential phonological posterior features,” Comput. Speech Lang., vol. 46, 2017, doi: 10.1016/j.csl.2017.06.004.

[21] T. Bent and R. F. Holt, “Representation of speech variability,” Wiley Interdiscip. Rev. Cogn. Sci., vol. 8, no. 4, 2017, doi: 10.1002/wcs.1434.

[22] D. F. Kleinschmidt, “Structure in talker variability: How much is there and how much can it help?,” Lang. Cogn. Neurosci., vol. 34, no. 1, 2019, doi: 10.1080/23273798.2018.1500698.

[23] B. McFee et al., “librosa: Audio and music signal analysis in Python,” in Proc. 14th Python Sci. Conf. (SciPy), 2015, doi: 10.25080/MAJORA-7B98E3ED-003.

[24] M. McAuliffe, M. Socolof, S. Mihuc, M. Wagner, and M. Sonderegger, “Montreal Forced Aligner: Trainable text-speech alignment using Kaldi,” in Proc. Interspeech 2017, 2017, pp. 498–502.

[25] Y. Sun, Y. Ming, X. Zhu, and Y. Li, “Out-of-distribution detection with deep nearest neighbors,” in Proc. 39th Int. Conf. Mach. Learn. (ICML), 2022. [Online]. Available: https://proceedings.mlr.press/v162/sun22d.html

[26] B. Schölkopf, J. C. Platt, J. Shawe-Taylor, A. J. Smola, and R. C. Williamson, “Estimating the support of a high-dimensional distribution,” Neural Comput., vol. 13, no. 7, pp. 1443–1471, 2001, doi: 10.1162/089976601750264965.

[27] M. Shahin and B. Ahmed, “Anomaly detection based pronunciation verification approach using speech attribute features,” Speech Commun., vol. 111, 2019, doi: 10.1016/j.specom.2019.06.003.

[28] K. Tang, R. Wayland, F. Wang, S. Vellozzi, R. Sengupta, and L. Altmann, “From sonority hierarchy to posterior probability as a measure of lenition: The case of Spanish stops,” J. Acoust. Soc. Amer., vol. 153, no. 2, 2023, doi: 10.1121/10.0017247.

[29] J. R. Duffy, E. A. Strand, H. Clark, M. Machulda, J. L. Whitwell, and K. A. Josephs, “Primary progressive apraxia of speech: Clinical features and acoustic and neurologic correlates,” Amer. J. Speech-Lang. Pathol., vol. 24, no. 2, 2015, doi: 10.1044/2015AJSLP-14-0174.

[30] X. Robin et al., “pROC: An open-source package for R and S+ to analyze and compare ROC curves,” BMC Bioinf., vol. 12, no. 1, 2011, doi: 10.1186/1471-2105-12-77.

[31] R: A Language and Environment for Statistical Computing. R Foundation for Statistical Computing, Vienna, Austria, 2025. [Online]. Available: https://www.R-project.org/

[32] D. Bates, M. Mächler, B. Bolker, and S. Walker, “Fitting linear mixed-effects models using lme4,” J. Stat. Softw., vol. 67, no. 1, 2015, doi: 10.18637/jss.v067.i01.

[33] A. Kuznetsova, P. B. Brockhoff, and R. H. B. Christensen, “lmerTest package: Tests in linear mixed effects models,” J. Stat. Softw., vol. 82, no. 13, 2017, doi: 10.18637/jss.v082.i13.

[34] emmeans: Estimated Marginal Means, aka Least-Squares Means. R Foundation for Statistical Computing, 2025. [Online]. Available: https://rvlenth.github.io/emmeans/

[35] C. Bhat and H. Strik, “Speech technology for automatic recognition and assessment of dysarthric speech: An overview,” J. Speech, Lang., Hear. Res., vol. 68, no. 2, 2025, doi: 10.1044/2024JSLHR-23-00740.

[36] W. Xue, R. van Hout, C. Cucchiarini, and H. Strik, “Assessing speech intelligibility of pathological speech in sentences and word lists: The contribution of phoneme-level measures,” J. Commun. Disord., vol. 102, 2023, doi: 10.1016/j.jcomdis.2023.106301.

[37] G. Van Nuffelen, C. Middag, M. De Bodt, and J.-P. Martens, “Speech technology-based assessment of phoneme intelligibility in dysarthria,” Int. J. Lang. Commun. Disord., vol. 44, no. 5, pp. 716–730, 2009, doi: 10.1080/13682820802342062.

[38] C. Middag et al., “Automated intelligibility assessment of pathological speech using phonological features,” EURASIP J. Adv. Signal Process., vol. 2009, no. 1, 2009, doi: 10.1155/2009/629030.

[39] M. P. Gelfer and Q. E. Bennett, “Speaking fundamental frequency and vowel formant frequencies: Effects on perception of gender,” J. Voice, vol. 27, no. 5, 2013, doi: 10.1016/j.jvoice.2012.11.008.

[40] V. G. Skuk and S. R. Schweinberger, “Influences of fundamental frequency, formant frequencies, aperiodicity, and spectrum level on the perception of voice gender,” J. Speech, Lang., Hear. Res., vol. 57, no. 1, 2014, doi: 10.1044/1092-4388(2013/12-0314).

